# Admission Predictors of In-Hospital Mortality and the Combined Outcome of Death or Invasive Mechanical Ventilation in Patients with COVID-19 During the Pre-Vaccination Era: A Retrospective Cohort Study

**DOI:** 10.64898/2026.02.28.26347308

**Authors:** Anis Rassi, Vanessa Mahamed Rassi, José Vitor Rassi Garcia, Hélvio Martins Gervásio, Christiane Reis Kobal, Fabiano Macedo de Souza, Gabriela Ferreira de Oliveira Butrico, Eddy Patrícia Sanchez, Fábio Mahamed Rassi, Germana Peres Canedo, Verônica Raquel Pinto Cunha, Raimundo Nonato Diniz Rodrigues Filho, Antonio Fernando Carneiro, Gustavo Gabriel Rassi

## Abstract

**Background:** Reliable identification of early predictors of adverse outcomes was essential during the pre-vaccination phase of the COVID-19 pandemic. Few studies have comprehensively integrated clinical presentation, laboratory parameters including arterial blood gas analysis, and chest computed tomography (CT) findings within a single well-characterized cohort, particularly in underrepresented regions of Brazil.

**Methods:** This retrospective cohort study included 482 consecutive adults (median age 61 years [IQR 49–73]; 64.3% men) with RT-PCR–confirmed SARS-CoV-2 infection hospitalized at a tertiary cardiac center in Central-West Brazil between March 2020 and January 2021. Demographic, clinical, laboratory (including arterial blood gas analysis), and chest CT data obtained within 48 hours of admission were analyzed. Univariable logistic regression was performed for 76 variables. Multivariable models were constructed using an a priori variable selection strategy based on clinical relevance, representation of distinct pathophysiological domains, and adherence to events-per-variable principles. Complete-case analyses were performed without imputation.

**Results:** In-hospital mortality was 9.3% (45/482). Invasive mechanical ventilation was required in 74 patients (15.4%), with a mortality rate of 58.1% among those ventilated. In univariable analysis, 42 variables were associated with mortality (p < 0.05). In multivariable analysis (n = 438), five independent predictors of death were identified: age (adjusted OR 1.66 per 10 years; 95% CI 1.19–2.32; p = 0.003), arterial pH (adjusted OR 0.47 per 0.1-unit increase; 95% CI 0.25–0.89; p = 0.021), neutrophil-to-lymphocyte ratio (adjusted OR 1.30; 95% CI 1.18–1.44; p < 0.001), number of comorbidities (adjusted OR 1.59; 95% CI 1.25–2.02; p < 0.001), and serum creatinine (adjusted OR 1.37; 95% CI 1.05–1.77; p = 0.019). The model demonstrated good calibration (Hosmer–Lemeshow p > 0.05) and moderate-to-high explanatory power (Nagelkerke R² = 0.43). For the composite outcome of death or invasive mechanical ventilation (74 events; 15.4%), four predictors remained independently associated; serum creatinine showed a non-significant trend (p = 0.069). On chest CT (n = 424), analyzed descriptively and in univariable models only, pulmonary involvement > 50% was associated with increased odds of death (OR 2.87; 95% CI 1.42–5.79; p = 0.003).

**Conclusions:** Five admission variables representing distinct pathophysiological domains—age, arterial pH, neutrophil-to-lymphocyte ratio, comorbidity burden, and serum creatinine—were independently associated with in-hospital mortality in this pre-vaccination cohort. Arterial pH provided independent prognostic information beyond inflammatory and renal markers. These findings support early risk stratification using routinely available clinical data.

## Introduction

The emergence of SARS-CoV-2 in late 2019 triggered one of the most profound global health crises in modern medicine [1]. Following the first reported cases in Wuhan, China, in December 2019 [2], COVID-19 rapidly spread worldwide, reaching Brazil in February 2020 [3]. The period preceding the introduction of vaccination in January 2021 represents a distinct and historically relevant phase of the pandemic, during which the disease manifested without population immunity or vaccine-induced modulation of outcomes [4–7].

During this early phase, hospitals faced unprecedented pressure, and clinical decision-making relied largely on observational evidence. Although numerous studies investigated predictors of severity and mortality, many were limited by heterogeneous methodologies, incomplete datasets, or insufficient adjustment for confounding factors [8,9]. Few studies systematically integrated clinical presentation, laboratory biomarkers—including arterial blood gas analysis—and chest computed tomography (CT) findings within a single well-characterized cohort. In Brazil, most published data originated from large metropolitan centers in the South, Southeast, and Northeast regions [10], leaving the Central-West region substantially underrepresented despite its distinct demographic and healthcare characteristics.

Well-documented pre-vaccination cohorts provide a valuable baseline of natural disease behavior, free from the confounding effects of vaccination, viral evolution, and widespread therapeutic standardization. Such data offer important reference points for understanding the pathophysiology of severe respiratory infections and for informing preparedness strategies for future pandemics.

In this context, the present study analyzes a large cohort of consecutive patients hospitalized with laboratory-confirmed COVID-19 at a tertiary cardiac center in Central-West Brazil during the first pandemic wave. Our objectives were to comprehensively describe admission characteristics and to identify independent predictors of in-hospital mortality and of a composite outcome of death or invasive mechanical ventilation, while evaluating the descriptive and prognostic value of chest CT findings through univariable analysis.

## Materials and Methods

### Study design and setting

This was a retrospective, single-center observational cohort study conducted at Hospital do Coração Anis Rassi (HCAR), a tertiary cardiac referral center in Goiânia, Goiás, Central-West Brazil. The study period spanned 321 days, from March 13, 2020 (date of the first suspected COVID-19 case at the institution) to January 28, 2021, corresponding to the pre-vaccination phase of the COVID-19 pandemic in Brazil. The national vaccination campaign in Brazil began on January 17, 2021. The study was reported in accordance with the Strengthening the Reporting of Observational Studies in Epidemiology (STROBE) guidelines.

### Patient selection

All consecutive patients evaluated for suspected COVID-19 during the study period underwent reverse transcription polymerase chain reaction (RT-PCR) testing for SARS-CoV-2. A total of 4,500 patients were evaluated (approximately 14 per day), of whom 1,161 (25.8%) tested positive, 3,301 (73.4%) tested negative, and 38 (0.8%) had inconclusive results. Among the 1,161 patients with confirmed COVID-19, 497 (42.8%) required hospitalization, while 664 (57.2%) were managed as outpatients.

Patients were eligible for inclusion if they were aged 18 years or older, had laboratory-confirmed COVID-19 by RT-PCR, and required hospital admission based on clinical judgment and institutional criteria in effect at the time. Exclusion criteria included leaving against medical advice (n = 3), inter-hospital transfer before a definitive outcome could be ascertained (n = 6), and death not attributable to COVID-19 (n = 6). After applying these criteria, 482 hospitalized patients constituted the final study cohort.

Most patients (451; 93.6%) were admitted through the HCAR Emergency Department, 20 (4.1%) were transferred from other hospitals, 7 (1.5%) had nosocomial COVID-19 acquired during hospitalization for other conditions, and 4 (0.8%) were referred from outpatient clinics. Regarding the initial care setting, 247 patients (51.2%) were admitted to general wards, 142 (29.5%) were admitted directly to the intensive care unit (ICU), and 93 (19.3%) were initially admitted to general wards but subsequently transferred to the ICU due to clinical deterioration. The complete patient flow from enrollment through clinical outcomes is depicted in **Fig 1**.

**Fig 1.**
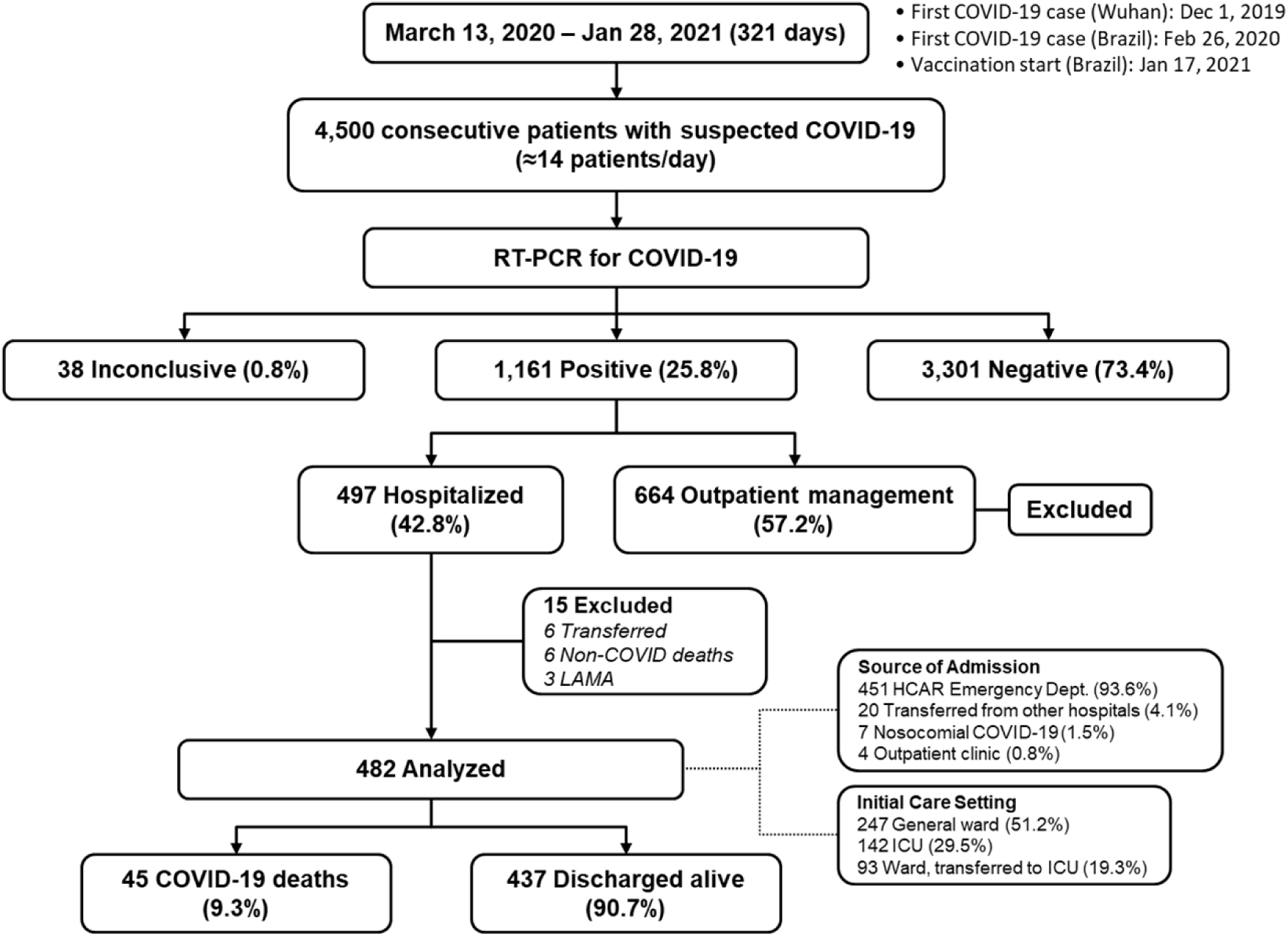
Flow diagram of patient enrollment, RT-PCR testing, and clinical outcomes among consecutive patients with suspected COVID-19 at Hospital do Coração Anis Rassi (HCAR), Goiânia, Brazil, March 2020 – January 2021. ICU: Intensive Care Unit; LAMA: leaving against medical advice.

### Outcomes

The primary outcome was in-hospital mortality attributable to COVID-19. The secondary outcome was a composite of in-hospital death or need for invasive mechanical ventilation. Outcomes were assessed during the index hospitalization only, with no post-discharge follow-up.

### Data collection

Data were extracted from electronic medical records by a trained team using a standardized electronic case report form, with consistency verification by two independent reviewers. Only variables available at hospital admission (during the first 48 hours) were considered for prognostic modeling.

Demographic and anthropometric data included age, sex, weight, height, body mass index (BMI), occupation (including whether the patient was a healthcare worker), and source of admission.

Vital signs at presentation included body temperature, heart rate, systolic and diastolic blood pressure, respiratory rate, and peripheral oxygen saturation (SpO₂) on room air.

Laboratory parameters obtained at admission included a comprehensive panel: complete blood count with differential (hemoglobin, hematocrit, leukocytes, lymphocytes, neutrophils, platelets), neutrophil-to-lymphocyte ratio (NLR), biochemical markers (urea, creatinine, sodium, potassium, total bilirubin, alanine aminotransferase [ALT], aspartate aminotransferase [AST], lactate dehydrogenase [LDH], C-reactive protein [CRP], ferritin, and troponin T), coagulation parameters (D-dimer, activated partial thromboplastin time [aPTT], prothrombin time, and international normalized ratio [INR]), and arterial blood gas analysis (pH, pO₂, pCO₂, bicarbonate, oxygen saturation, base excess, and lactic acid).

Pre-existing comorbidities were systematically recorded, including hypertension, type 2 diabetes mellitus, obesity (BMI ≥30 kg/m²), coronary artery disease, chronic obstructive pulmonary disease (COPD), chronic kidney disease (CKD), heart failure, malignancy, and smoking history.

Symptoms at presentation were documented, including dyspnea, fatigue, dry cough, fever, sore throat, headache, anosmia, ageusia, nausea/vomiting, diarrhea, and altered mental status (irritability or confusion).

Disease severity at admission was classified according to both World Health Organization (WHO) classification (non-severe, severe, and critical) [11] and the National Institutes of Health (NIH) classification (mild, moderate, severe, and critical) [12].

In-hospital data included therapeutic interventions (corticosteroids, convalescent plasma, colchicine, anticoagulation, remdesivir, and other agents), oxygen support modalities (nasal cannula, non-rebreather mask, high-flow nasal cannula, non-invasive and invasive mechanical ventilation, prone positioning), vasopressor use, neuromuscular blockade, tracheostomy, blood transfusion, and renal replacement therapy. Complications were recorded as they occurred, including acute respiratory distress syndrome (ARDS), bacterial pneumonia, acute kidney injury (AKI), septic shock, thromboembolic events, stroke, cardiac arrhythmias, and new-onset heart failure. Time from symptom onset to hospitalization was also recorded.

### Chest computed tomography

Chest CT was performed according to institutional protocols and was available for analysis in 424 patients. Scans were excluded if image quality was inadequate or if significant pre-existing pulmonary disease was present. All 424 chest CT scans were retrospectively reviewed by a single experienced thoracic radiologist who was blinded to clinical outcomes, using a standardized structured reporting protocol. This protocol encompassed predominant imaging patterns, distribution of lesions, ancillary findings, and a final classification regarding likelihood of COVID-19 pneumonia. This systematic approach ensured consistency in the assessment and eliminated interobserver variability. Pulmonary involvement was visually estimated and categorized into six groups by extent of parenchymal involvement: no pulmonary involvement, 1–10%, 11–25%, 26–50%, 51–75%, and > 75%. For analytical purposes, pulmonary involvement was also dichotomized as 1-50% versus > 50%. Given incomplete availability and potential collinearity with clinical markers of disease severity, chest CT variables were analyzed descriptively and in univariable models only and were not included in multivariable regression analyses.

### Ethical considerations

This retrospective study was approved by the Institutional Research Ethics Committee of Hospital do Coração Anis Rassi (CAAE 85626624.0.3001.5075). Given the retrospective nature of the study and the use of de-identified data extracted from electronic medical records, the requirement for individual informed consent was waived by the ethics committee. Data confidentiality was preserved throughout the study in accordance with applicable regulations.

### Statistical analysis

Continuous variables were tested for normality using the Kolmogorov–Smirnov test and are presented as median and interquartile range (IQR). Categorical variables are expressed as absolute numbers and percentages. Comparisons between groups were performed using the Mann–Whitney U test for continuous variables and the chi-square or Fisher’s exact test for categorical variables, as appropriate.

Univariable logistic regression analyses were initially performed to evaluate the association between each variable and the outcomes of interest. Variables with more than 10% missing data were excluded from multivariable analysis to minimize selection bias; these included troponin T (52.1% missing), lactic acid (64.1% missing), D-dimer (37.3% missing), and total bilirubin (41.3% missing), although these variables were analyzed in univariable models. Variables exhibiting complete or quasi-complete separation (i.e., 0% or 100% occurrence in one outcome group, precluding odds ratio estimation) were also excluded from multivariable modeling. Furthermore, variables representing in-hospital treatments and complications were not included in multivariable analysis, as these represent consequences of disease progression rather than baseline risk factors and could introduce reverse causation bias.

Variables considered clinically relevant and those showing statistical significance in univariable analysis were selected for multivariable modeling. Multivariable analysis was conducted using binary logistic regression with the Enter method, in which all selected variables were simultaneously included in the model. This approach was chosen to preserve clinical interpretability and avoid data-driven variable selection. To reduce the risk of overfitting, the number of variables included in each model was limited according to the number of observed events. Collinearity among highly correlated variables was assessed, and when present, only the most clinically relevant variable was retained. Variables were selected to represent distinct pathophysiological domains — demographics, comorbidity burden, inflammatory response, metabolic status, and organ dysfunction — ensuring complementary prognostic information. Patients with incomplete data for any covariate were excluded from each model (complete-case analysis).

Chest CT variables were not included in multivariable models due to incomplete availability and potential collinearity with clinical markers of disease severity, but were analyzed descriptively and in univariable models.

Model calibration was assessed using the Hosmer–Lemeshow goodness-of-fit test, and model performance was evaluated using Cox & Snell R² and Nagelkerke’s R². Results are reported as odds ratios (OR) with 95% confidence intervals (CI), and a two-sided p-value < 0.05 was considered statistically significant. All analyses were performed using SPSS version 15.0 (SPSS Inc., Chicago, IL, USA).

## Results

### Study population and baseline characteristics

A total of 482 patients with RT-PCR–confirmed SARS-CoV-2 infection were included in the study. The overall in-hospital mortality rate was 9.3% (45/482). The median age of the cohort was 61 years (IQR 49–73; range 18–99), and 310 patients (64.3%) were male. The median BMI was 27.4 kg/m² (IQR 25.0–30.4). Healthcare professionals accounted for 17.8% of admissions and had significantly lower odds of death (OR 0.20; 95% CI 0.05–0.82; p = 0.026).

Non-survivors were significantly older than survivors (median 74 vs. 59 years; p < 0.001), with each 10-year increase in age associated with a twofold increase in the odds of death (OR 2.00; 95% CI 1.56–2.57). Sex and BMI did not differ significantly between groups.

### Comorbidities

At least one comorbidity was present in 384 patients (79.7%), and the burden was significantly higher among non-survivors (93.3% vs. 78.3%; p = 0.026), who had a median of 3 comorbidities (IQR 2–4) compared with 1 (IQR 1–3) in survivors (p < 0.001). The most prevalent comorbidities in the overall cohort were hypertension (55.2%), type 2 diabetes mellitus (27.4%), and obesity (27.2%) **(S1 Fig).** Among these, hypertension (OR 2.14; 95% CI 1.09–4.18) and diabetes (OR 2.10; 95% CI 1.12–3.93) were significantly associated with mortality, whereas obesity was not (p = 0.787).

Heart disease (excluding hypertension) was present in 18.0% of patients and was strongly associated with death (OR 2.54; 95% CI 1.30–4.95; p = 0.006), with coronary artery disease (10.2%), heart failure (3.1%), and cardiac device carriers (1.7%) all contributing to this excess risk. Chronic kidney disease — both non-dialytic (17.8% vs. 2.5%; OR 8.37) and dialytic (8.9% vs. 2.1%; OR 4.64) — showed among the strongest associations with mortality. Other comorbidities significantly linked to death included COPD (OR 4.28; p = 0.001), malignant neoplasia (OR 4.05; p = 0.006), former smoking (OR 4.50; p < 0.001), bronchial asthma (OR 3.46; p = 0.039), and moderate-to-severe liver disease (OR 10.12; p = 0.022). Detailed demographic and comorbidity data are presented in **Table 1**.

**Table 1.** Baseline demographics, comorbidities, symptoms, and disease severity according to in-hospital mortality.

| Variable | N | Total (N=482) | Death (n=45) | Survivor (n=437) | OR (95% CI) | p |
| --- | --- | --- | --- | --- | --- | --- |
| **Demographics** |  |  |  |  |  |  |
| Age (per 10 years) | 482 | 61 (49–73) | 74 (69–80) | 59 (48–71) | 1.999 (1.556–2.568) | **<0.001** |
| Male sex | 482 | 310 (64.3) | 29 (64.4) | 281 (64.3) | 1.006 (0.530–1.910) | 0.985 |
| BMI, kg/m<sup>2</sup> | 470 | 27.4 (25.0–30.4) | 27.1 (23.8–29.4) | 27.4 (25.2–30.5) | 0.946 (0.879–1.018) | 0.139 |
| Healthcare professional | 482 | 86 (17.8) | 2 (4.4) | 84 (19.2) | 0.195 (0.046–0.823) | **0.026** |
| **Comorbidities** |  |  |  |  |  |  |
| Any comorbidity | 482 | 384 (79.7) | 42 (93.3) | 342 (78.3) | 3.889 (1.179–12.823) | **0.026** |
| Number of comorbidities | 482 | 2 (1–3) | 3 (2–4) | 1 (1–3) | 1.733 (1.436–2.090) | **<0.001** |
| Hypertension | 482 | 266 (55.2) | 32 (71.1) | 234 (53.5) | 2.135 (1.091–4.179) | **0.027** |
| Diabetes mellitus type 2 | 482 | 132 (27.4) | 19 (42.2) | 113 (25.9) | 2.095 (1.117–3.931) | **0.021** |
| Obesity (BMI ≥30) | 482 | 131 (27.2) | 13 (28.9) | 118 (27.0) | 1.098 (0.557–2.164) | 0.787 |
| Heart disease* | 482 | 87 (18.0) | 15 (33.3) | 72 (16.5) | 2.535 (1.298–4.950) | **0.006** |
| Coronary artery disease | 482 | 49 (10.2) | 9 (20.0) | 40 (9.2) | 2.481 (1.115–5.520) | **0.026** |
| Heart failure | 482 | 15 (3.1) | 4 (8.9) | 11 (2.5) | 3.778 (1.151–12.400) | **0.028** |
| Pacemaker/ICD/CRT | 482 | 8 (1.7) | 3 (6.7) | 5 (1.1) | 6.171 (1.425–26.734) | **0.015** |
| Chagas heart disease | 482 | 6 (1.2) | 1 (2.2) | 5 (1.1) | 1.964 (0.224–17.186) | 0.542 |
| Valvular heart disease | 482 | 12 (2.5) | 2 (4.4) | 10 (2.3) | 1.986 (0.421–9.359) | 0.386 |
| Non-dialytic CKD | 482 | 19 (3.9) | 8 (17.8) | 11 (2.5) | 8.373 (3.172–22.104) | **<0.001** |
| Dialytic CKD | 482 | 13 (2.7) | 4 (8.9) | 9 (2.1) | 4.640 (1.369–15.725) | **0.014** |
| COPD | 482 | 29 (6.0) | 8 (17.8) | 21 (4.8) | 4.283 (1.775–10.337) | **0.001** |
| Bronchial asthma | 482 | 16 (3.3) | 4 (8.9) | 12 (2.7) | 3.455 (1.066–11.201) | **0.039** |
| Malignant neoplasia | 482 | 22 (4.6) | 6 (13.3) | 16 (3.7) | 4.048 (1.498–10.938) | **0.006** |
| Liver disease (mod.–severe) | 482 | 4 (0.8) | 2 (4.4) | 2 (0.5) | 10.12 (1.390–73.626) | **0.022** |
| Former smoking | 482 | 32 (6.6) | 9 (20.0) | 23 (5.3) | 4.500 (1.938–14.450) | **<0.001** |
| Current smoking | 482 | 10 (2.1) | 2 (4.4) | 8 (1.8) | 2.494 (0.513–12.120) | 0.257 |
| **Symptoms at admission** |  |  |  |  |  |  |
| Time since onset, days | 451 | 8 (7–10) | 7 (4–9) | 9 (7–10) | 0.853 (0.764–0.951) | **0.004** |
| Shortness of breath | 482 | 300 (62.2) | 33 (73.3) | 267 (61.1) | 1.751 (0.880–3.484) | 0.111 |
| Fatigue | 482 | 267 (55.4) | 20 (44.4) | 247 (56.5) | 0.615 (0.332–1.141) | 0.123 |
| Dry cough | 482 | 261 (54.1) | 28 (62.2) | 233 (53.3) | 1.442 (0.767–2.711) | 0.256 |
| Fever | 482 | 257 (53.3) | 21 (46.7) | 236 (54.0) | 0.745 (0.403–1.379) | 0.349 |
| Muscle pain | 482 | 173 (35.9) | 11 (24.4) | 162 (37.1) | 0.549 (0.271–1.114) | 0.549 |
| Headache | 482 | 109 (22.6) | 6 (13.3) | 103 (23.6) | 0.449 (0.205–1.212) | 0.125 |
| Diarrhea | 482 | 95 (19.7) | 9 (20.0) | 86 (19.7) | 1.020 (0.474–2.198) | 0.959 |
| Nausea/vomiting | 482 | 75 (15.6) | 7 (15.6) | 68 (15.6) | 1.000 (0.429–2.331) | 0.999 |
| Chest pain | 482 | 74 (15.4) | 5 (11.1) | 69 (15.8) | 0.667 (0.254–1.749) | 0.410 |
| Loss of smell | 482 | 51 (10.6) | 3 (6.7) | 48 (11.0) | 0.579 (0.173–1.939) | 0.376 |
| Loss of taste | 482 | 28 (5.8) | 3 (6.7) | 25 (5.7) | 1.177 (0.341–4.063) | 0.796 |
| Sore throat | 482 | 46 (9.5) | 1 (2.2) | 45 (10.3) | 0.198 (0.027–1.472) | 0.114 |
| Irritability/confusion | 482 | 19 (3.9) | 5 (11.1) | 14 (3.2) | 3.777 (1.294–11.026) | **0.015** |
| **Disease severity at admission** |  |  |  |  |  |  |
| Classification 1† | 482 |  |  |  |  |  |
| Non-severe |  | 321 (66.6) | 11 (24.4) | 310 (70.9) |  |  |
| Severe |  | 136 (28.2) | 23 (51.1) | 113 (25.9) |  |  |
| Critical |  | 25 (5.2) | 11 (24.4) | 14 (3.2) |  |  |
| Severe/critical vs non-severe | 482 | 161 (33.4) | 34 (75.6) | 127 (29.1) | 7.545 (3.707–15.354) | **<0.001** |
| Classification 2‡ | 482 |  |  |  |  |  |
| Mild |  | 35 (7.3) | 0 (0.0) | 35 (8.0) |  |  |
| Moderate |  | 240 (49.8) | 10 (22.2) | 230 (52.6) |  |  |
| Severe |  | 187 (38.8) | 24 (53.3) | 163 (37.3) |  |  |
| Critical |  | 20 (4.1) | 11 (24.4) | 9 (2.1) |  |  |
| Severe/critical vs mild/mod. | 482 | 207 (42.9) | 35 (77.8) | 172 (39.4) | 5.392 (2.603–11.173) | **<0.001** |

Data are presented as median (IQR) for continuous variables or n (%) for categorical variables. OR, odds ratio from univariate logistic regression (per unit increase unless otherwise specified in parentheses); CI, confidence interval; BMI, body mass index; CKD, chronic kidney disease; COPD, chronic obstructive pulmonary disease; ICD, implantable cardioverter-defibrillator; CRT, cardiac resynchronization therapy. \* Excluding hypertension. † WHO 3-category classification. ‡ NIH 4-category classification.

### Symptoms and disease severity

The median time from symptom onset to hospital admission was 8 days (IQR 7–10) **(S2 Fig)** and was significantly shorter in non-survivors (7 vs. 9 days; OR 0.85 per day; p = 0.004) **(Table 1).** The most common presenting symptoms were shortness of breath (62.2%), fatigue (55.4%), dry cough (54.1%), and fever (53.3%) **(S3 Fig).** Most individual symptoms did not differ significantly between groups; however, irritability or confusion at presentation was nearly four times more common among non-survivors (11.1% vs. 3.2%; OR 3.78; p = 0.015) **(Table 1).**

According to the WHO classification, 66.6% of patients were classified as non-severe at admission, 28.2% as severe, and 5.2% as critical. By the NIH 4-category classification, 7.3% were mild, 49.8% moderate, 38.8% severe, and 4.1% critical **(S4 Fig).** Both severity systems strongly predicted mortality: severe or critical disease at admission was associated with an OR of 7.55 (95% CI 3.71–15.35; p < 0.001) by the WHO classification and 5.39 (95% CI 2.60–11.17; p < 0.001) by the NIH classification **(Table 1).**

### Vital signs and laboratory findings

Admission vital signs revealed significantly lower oxygen saturation on ambient air among non-survivors (median 88.5% vs. 94%; OR 0.79 per percentage point; p < 0.001), higher respiratory rate (19 vs. 18 cpm; OR 1.17; p < 0.001), and higher systolic blood pressure (134 vs. 126 mmHg; OR 1.02; p = 0.029). Heart rate, temperature, and diastolic blood pressure did not differ significantly between groups.

Among hematological parameters, non-survivors had lower hemoglobin (12.4 vs. 13.8 g/dL; OR 0.74; p < 0.001) and markedly elevated neutrophil-to-lymphocyte ratio (NLR; 9.4 vs. 4.8; OR 1.26; p < 0.001), reflecting both absolute lymphopenia (732 vs. 1107/mL; p < 0.001) and a shift toward neutrophilic inflammation. Biochemical markers of organ dysfunction — urea (52 vs. 30 mg/dL), creatinine (1.35 vs. 1.01 mg/dL), LDH (648 vs. 564 U/L), and CRP (129.1 vs. 62.7 mg/L) — were all significantly elevated in non-survivors. D-dimer levels were approximately twofold higher in non-survivors (0.75 vs. 0.4 mg/mL; OR 2.02; p < 0.001). Ferritin and troponin T levels did not differ significantly between groups, although troponin T was available for only 231 patients (47.9%).

Arterial blood gas analysis showed that non-survivors had lower pH (7.40 vs. 7.42; OR 0.29 per 0.1 unit; p < 0.001), lower bicarbonate (21.4 vs. 22.1 mmol/L; OR 0.85; p = 0.010), and more negative base excess (−2.9 vs. −1.3 mmol/L; OR 0.74; p < 0.001), indicating a tendency toward metabolic acidosis. Vital signs and complete laboratory data are shown in **Table 2**.

**Table 2.**
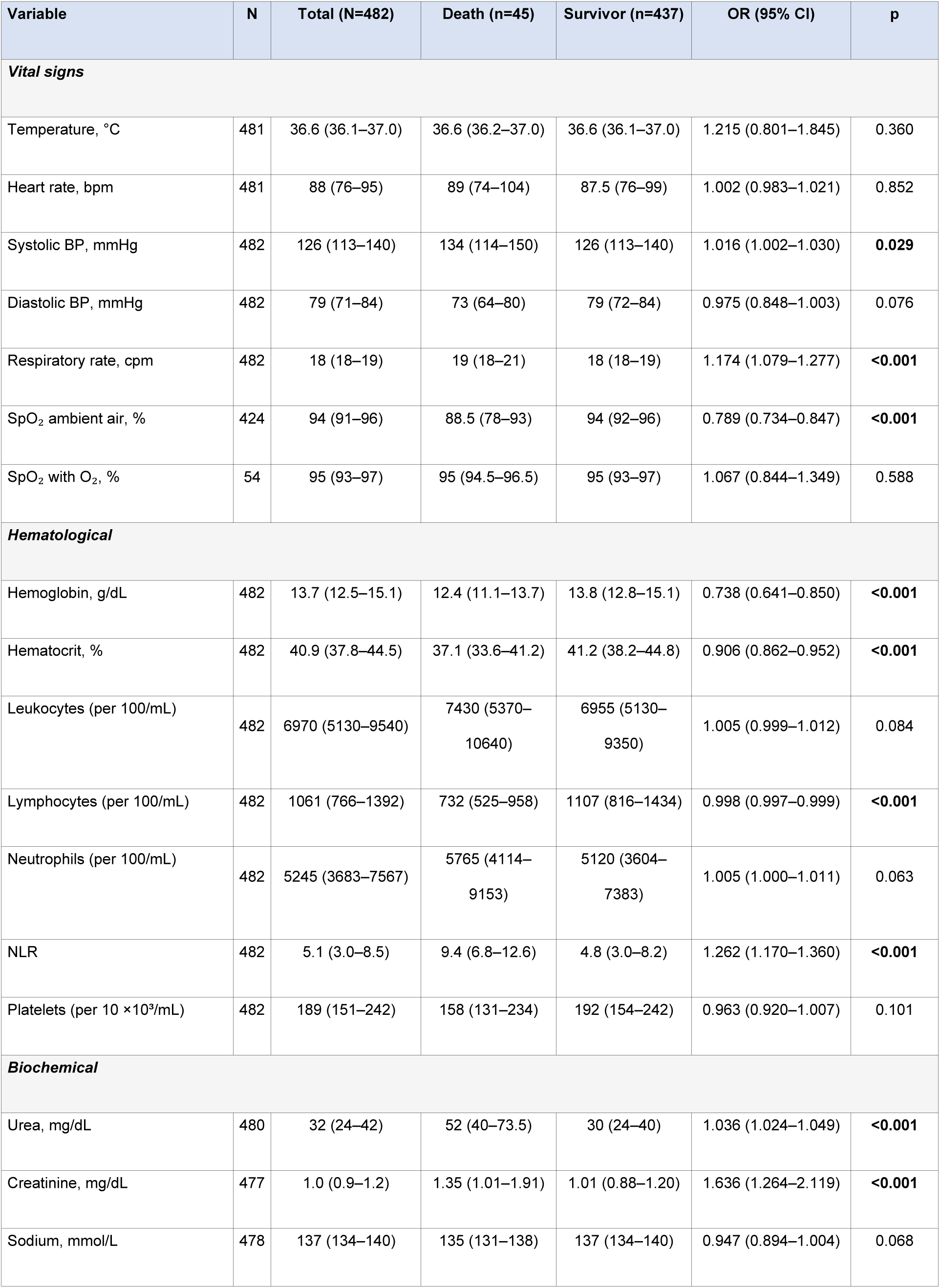
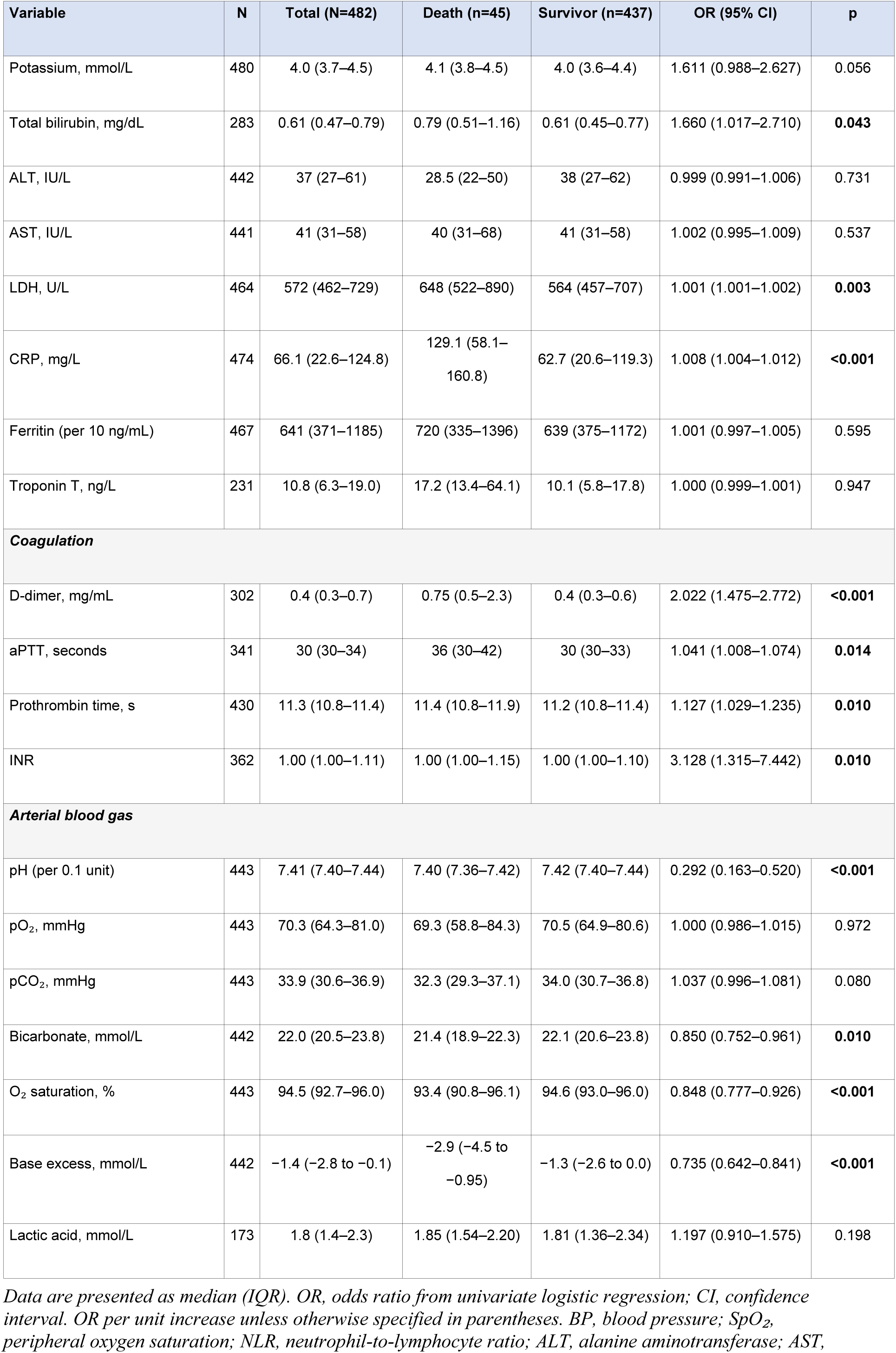
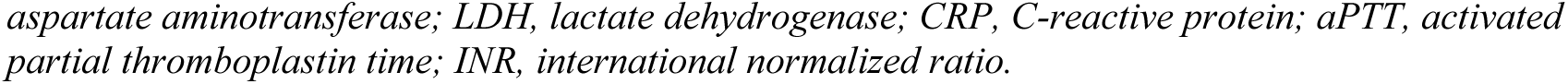
Admission vital signs and laboratory parameters according to in-hospital mortality.

### Chest CT findings

Chest CT was available for analysis in 424 of 482 patients (88.0%), including 35 of 45 non-survivors (77.8%) and 389 of 437 survivors (89.0%). Among these, 406 had identifiable pulmonary involvement, while 18 showed no parenchymal abnormalities.

The predominant radiological pattern was ground-glass opacity, observed in 325 patients (76.6%), followed by mixed ground-glass and consolidation in 72 (17.0%) and predominant consolidation in 27 (6.4%). Pulmonary involvement was bilateral in 340 patients (80.1%) and multifocal in 351 (82.8%), with a peripheral distribution in 248 (58.6%). Ancillary findings included pleural effusion in 80 patients (18.9%), bronchial dilatation in 53 (12.5%), reverse halo sign in 39 (9.2%), and septal thickening in 28 (6.6%). The overall CT pattern was classified as typical for COVID-19 in 336 patients (79.3%). Individual morphological patterns and distribution features did not differ significantly between survivors and non-survivors (data not shown).

Among the 406 patients with pulmonary involvement on CT, extension exceeding 50% was observed in 118 (29.1%) and was significantly more frequent in non-survivors (51.4% vs. 27.0%; OR 2.87; 95% CI 1.42–5.79; p = 0.003), indicating that global disease burden, rather than specific radiological patterns, was the principal CT-derived marker of severity. Complete CT data are presented in **Table 3**.

**Table 3.**
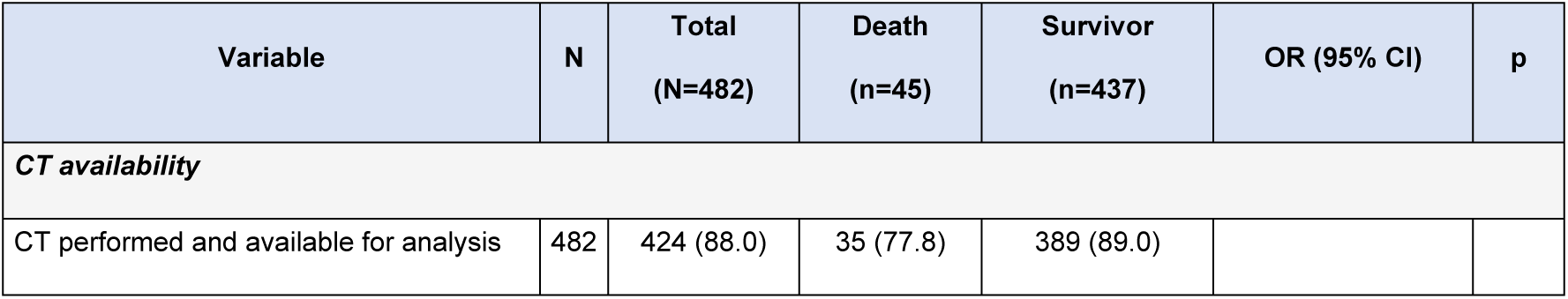
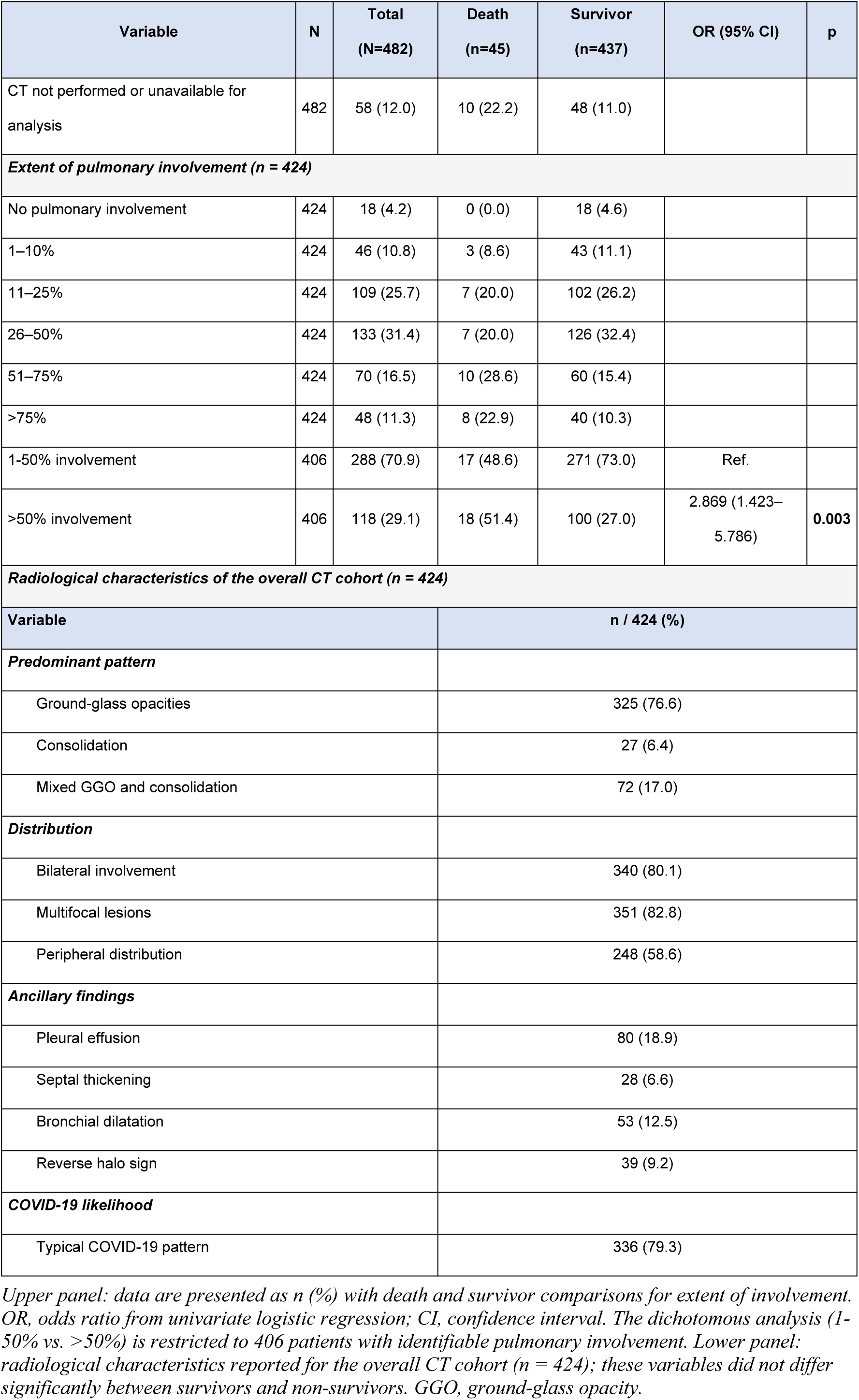
Chest computed tomography findings at hospital admission.

Representative CT images illustrating the spectrum of pulmonary involvement according to the European Society of Radiology/European Society of Thoracic Imaging (ESR/ESTI) visual quantification system [13] are presented in **Fig 2**.

**Fig 2.**
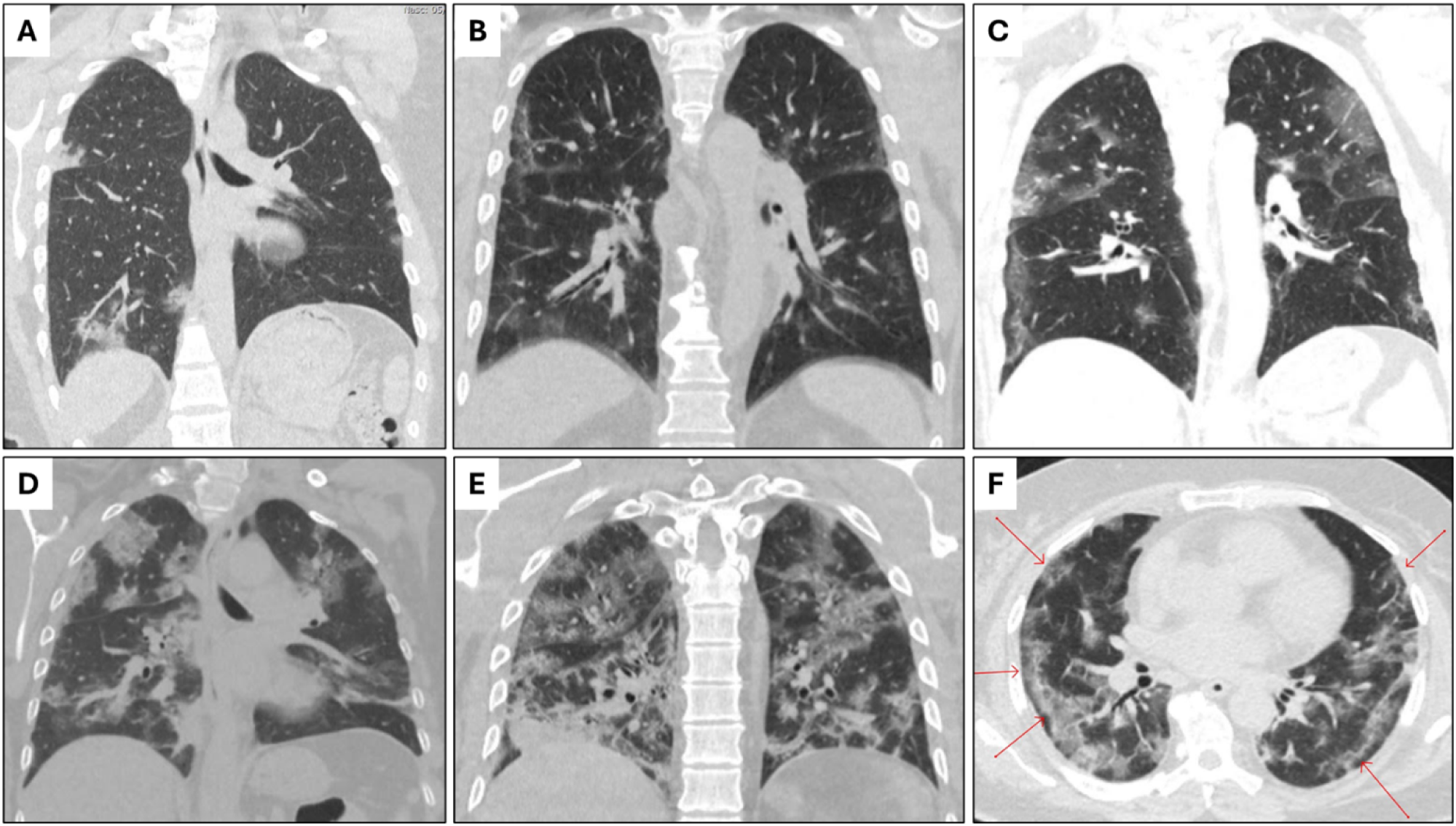
Representative chest CT images illustrating the spectrum of COVID-19 pulmonary involvement. Coronal (A–E) and axial (F) chest CT images from five different patients demonstrating increasing extent of ground-glass opacity (GGO), classified according to the European Society of Radiology/European Society of Thoracic Imaging (ESR/ESTI) visual quantification system [13]. (A) Minimal involvement (1–10%): sparse peripheral GGO in the right lower lobe. (B) Moderate involvement (11–25%): bilateral peripheral GGO predominantly affecting the lower lobes. (C) Important involvement (26–50%): bilateral, multifocal GGO with peripheral predominance involving all lobes. (D) Severe involvement (51–75%): extensive bilateral GGO with areas of consolidation and minimal residual aerated parenchyma. (E) Critical involvement (>75%): near-complete bilateral opacification with diffuse GGO and consolidation. (F) Axial view highlighting the hallmark CT features of COVID-19 pneumonia: ground-glass opacities with bilateral, multifocal, and peripheral distribution (red arrows). All images were obtained at the time of hospital admission.

### In-hospital complications

Complications occurred in 428 patients (88.8%) and were universal among non-survivors (100%) **(S1 Table).** ARDS was the most impactful complication, occurring in 77.8% of non-survivors versus 11.4% of survivors (OR 27.09; p < 0.001). Septic shock developed in 44.4% of non-survivors (OR 28.33; p < 0.001). Acute kidney injury was present in 60.0% of those who died, compared with only 2.3% of survivors (OR 64.05; p < 0.001), and nearly half of non-survivors required new dialysis (48.9% vs. 1.6%; OR 58.76; p < 0.001). Stroke occurred in 6.7% of non-survivors versus 0.2% of survivors (OR 31.14; p = 0.003). Bacterial pneumonia superimposed on viral pneumonia was documented in 53.3% of non-survivors compared with 24.0% of survivors (OR 3.61; p < 0.001). Complete complication data are provided in **S5 Fig.**

### Admission location, medications, oxygen therapy, and treatment interventions

Nearly half of patients (48.8%) required ICU admission at some point during hospitalization, and ICU admission was almost universal among non-survivors (97.8%; OR 56.67; p < 0.001). The median length of stay was 8 days (IQR 5–12; range 1–95) overall **(S6 Fig)** and was significantly longer for non-survivors (13 vs. 8 days; p < 0.001).

Corticosteroids were administered to 80.3% of patients, without significant difference between groups (p = 0.464). Convalescent plasma was used in 27.0% and anticoagulation in 95.2%, with neither showing a significant association with mortality **(S7 Fig).** Remdesivir was used in only 8 patients (1.7%). Colchicine use was significantly lower among non-survivors (4.4% vs. 25.9%; OR 0.13; p = 0.006), a finding that should be interpreted cautiously given its observational nature and the potential for confounding by indication.

Nearly all patients received oxygen therapy (94.2%) **(S8 Fig),** and the use of invasive mechanical ventilation was the strongest treatment-related predictor of death (95.6% of non-survivors vs. 7.1% of survivors; OR 281.60; p < 0.001). Non-invasive ventilation was employed in 40.0% of the cohort and was more frequent among non-survivors (64.4% vs. 37.5%; OR 3.02; p = 0.001), as were prone positioning (46.7% vs. 5.3%; OR 15.75; p < 0.001) and neuromuscular blockade (53.3% vs. 3.2%; OR 34.53; p < 0.001). Vasopressors were used in 97.8% of non-survivors (OR 642.70; p < 0.001). Admission location, medications, oxygen therapy, and treatment data are detailed in **S2 Table, S7 Fig, and S8 Fig.**

### Multivariable analysis

Among the 42 variables with p < 0.05 in univariable analysis **(S9 Fig),** five were selected for multivariable modeling based on clinical relevance, representation of distinct pathophysiological domains, and adherence to the rule of approximately 10 events per predictor variable. After excluding 44 patients with incomplete data for one or more covariates, 438 patients were included in the final models (complete-case analysis).

In the primary model for in-hospital death **(Table 4),** all five covariates were independently associated with mortality. Older age remained the strongest demographic predictor (adjusted OR 1.66 per 10 years; 95% CI 1.19–2.32; p = 0.003). Higher neutrophil-to-lymphocyte ratio (adjusted OR 1.30 per unit; 95% CI 1.18–1.44; p < 0.001) and a greater number of comorbidities (adjusted OR 1.59 per unit; 95% CI 1.25–2.02; p < 0.001) were also independently associated with death. Lower arterial pH at admission was a strong independent predictor, with each 0.1-unit increase conferring a 53% reduction in the odds of death (adjusted OR 0.47; 95% CI 0.25–0.89; p = 0.021).

**Table 4.**
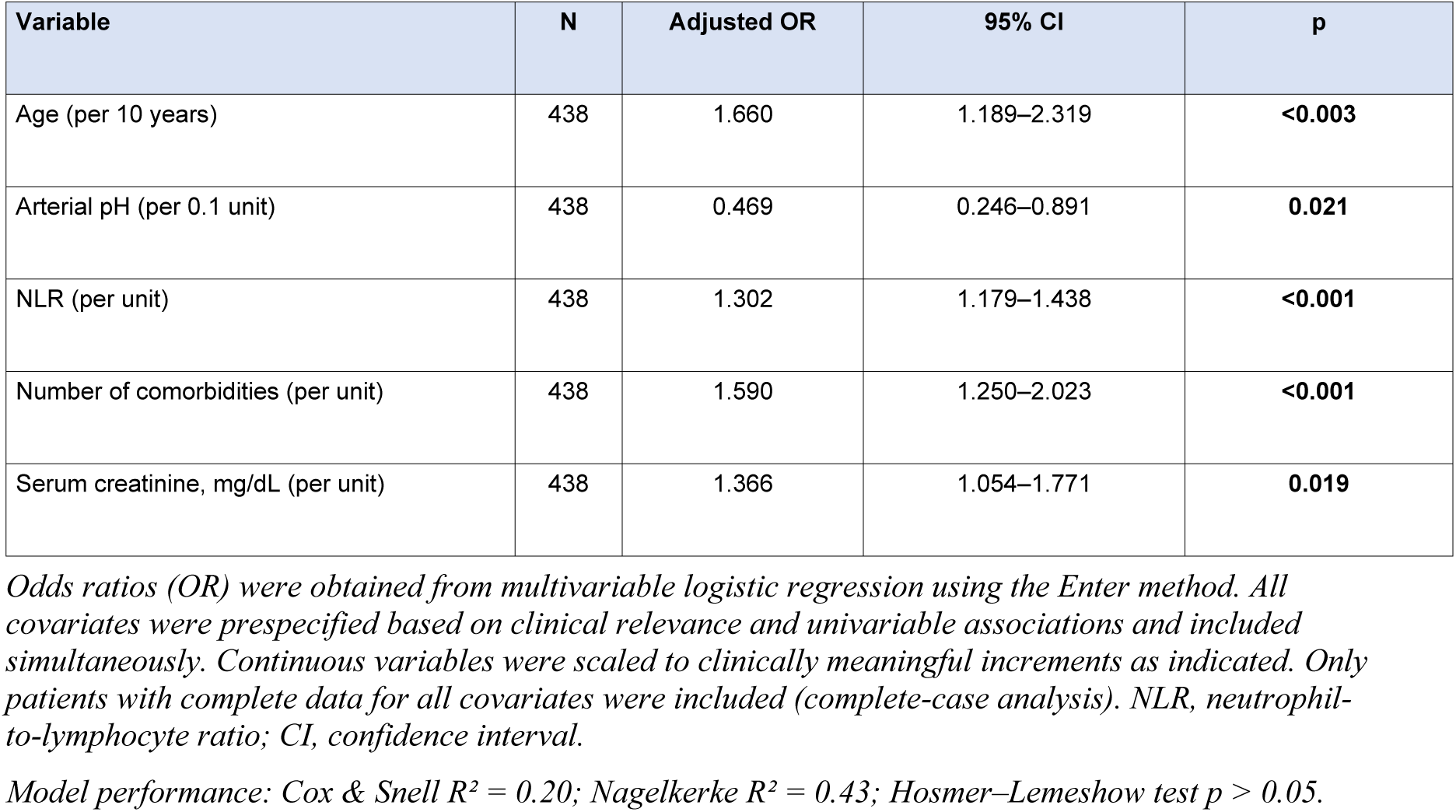
Multivariable logistic regression analysis for in-hospital death (primary outcome).

Baseline renal dysfunction, assessed by serum creatinine, retained independent significance (adjusted OR 1.37; 95% CI 1.05–1.77; p = 0.019). The model demonstrated good calibration (Hosmer–Lemeshow p > 0.05) and moderate-to-high explanatory power (Nagelkerke R² = 0.43).

For the secondary composite outcome of death or invasive mechanical ventilation (n = 74 events; 15.4%), the same five covariates were entered to allow direct comparison **(Table 5).** Age (adjusted OR 1.36 per 10 years; p = 0.009), arterial pH (adjusted OR 0.52 per 0.1 unit; p = 0.019), NLR (adjusted OR 1.27; p < 0.001), and number of comorbidities (adjusted OR 1.31; p = 0.006) remained independently associated with the composite outcome. In contrast, serum creatinine did not retain statistical significance (adjusted OR 1.26; 95% CI 0.98–1.61; p = 0.069), although a trend toward increased risk was observed. Model performance was adequate but lower than for the primary outcome (Nagelkerke R² = 0.34), reflecting the broader clinical heterogeneity of the composite endpoint.

**Table 5.** Multivariable logistic regression analysis for death or invasive mechanical ventilation (secondary outcome).

| Variable | N | Adjusted OR | 95% CI | p |
| --- | --- | --- | --- | --- |
| Age (per 10 years) | 438 | 1.358 | 1.078–1.710 | **0.009** |
| Arterial pH (per 0.1 unit) | 438 | 0.517 | 0.297–0.899 | **0.019** |
| NLR (per unit) | 438 | 1.272 | 1.179–1.373 | **<0.001** |
| Number of comorbidities (per unit) | 438 | 1.314 | 1.081–1.597 | **0.006** |
| Serum creatinine, mg/dL (per unit) | 438 | 1.257 | 0.983–1.608 | 0.069 |

*Odds ratios (OR) were obtained from multivariable logistic regression using the Enter method. All covariates were prespecified based on clinical relevance and univariable associations and included simultaneously. Continuous variables were scaled to clinically meaningful increments as indicated. Only patients with complete data for all covariates were included (complete-case analysis). NLR, neutrophil-to-lymphocyte ratio; CI, confidence interval.*
*Model performance: Cox & Snell $R^2 = 0.20$ ; Nagelkerke $R^2 = 0.34$ ; Hosmer–Lemeshow test $p > 0.05$ .*

## Discussion

This study provides a comprehensive evaluation of demographic, clinical, laboratory, and radiological predictors of in-hospital mortality in 482 consecutive patients hospitalized with RT-PCR–confirmed COVID-19 during the pre-vaccination phase of the pandemic in Central-West Brazil. The overall in-hospital mortality rate was 9.3%, and five independent predictors of death were identified: older age, lower arterial pH, higher NLR, greater number of comorbidities, and elevated serum creatinine. These variables represent distinct pathophysiological domains—demographics, acid–base homeostasis, systemic inflammation, comorbidity burden, and renal function—and collectively achieved moderate-to-high explanatory power (Nagelkerke R² = 0.43), a noteworthy result given that only five covariates were included to respect the constraint of approximately 10 events per predictor variable. The observed mortality rate is consistent with or lower than rates reported in other pre-vaccination cohorts. Early reports from Wuhan described mortality rates of 28% among hospitalized patients [5], while Grasselli et al. reported 26% ICU mortality in Lombardy [6]. In New York City, mortality among hospitalized patients reached 21% during the initial surge [7]. In Brazil, national registry data reported substantially higher mortality rates, reaching 41% in hospitalized patients [10]. Differences in patient selection, healthcare infrastructure, and disease severity at admission likely account for part of this variation. In our cohort, 66.6% of patients were classified as non-severe at admission, and systematic early evaluation—including arterial blood gas analysis in 92% and chest CT in 88%—may have contributed to risk stratification and management.

Age was the strongest independent predictor of mortality, with a 66% increase in risk per decade (adjusted OR 1.66; 95% CI 1.19–2.32; p < 0.003). This finding aligns with multiple international cohorts [5,6,14,15] and reflects mechanisms including immunosenescence, reduced physiological reserve, and increased susceptibility to thromboinflammatory responses [16,17]. The attenuation—but persistence—of the age effect in the composite outcome model suggests that while age strongly predicts death, progression to invasive mechanical ventilation may be mediated by additional physiological factors.

The cumulative burden of comorbidities, rather than individual diseases, independently predicted mortality (adjusted OR 1.59 per additional comorbidity; 95% CI 1.25–2.02). In univariable analyses, chronic kidney disease, liver disease, cardiac device carriers, COPD, malignancy, heart failure, coronary artery disease, hypertension, type 2 diabetes mellitus, and former smoking showed strong associations with mortality. Using the total number of comorbidities avoided collinearity and provided a parsimonious measure of baseline vulnerability. This approach is consistent with prior evidence demonstrating the additive impact of multimorbidity on COVID-19 outcomes [9,14].

The neutrophil-to-lymphocyte ratio emerged as a strong independent predictor (adjusted OR 1.30; 95% CI 1.18–1.44; p < 0.001), reflecting immune dysregulation as a central mechanism of severe disease. Elevated NLR integrates neutrophilia driven by systemic inflammation and lymphopenia associated with impaired adaptive immunity. Meta-analyses have confirmed its prognostic value across diverse populations [18,19], and its availability from routine blood testing enhances its clinical applicability.

Arterial pH was independently associated with mortality (adjusted OR 0.47 per 0.1-unit increase; 95% CI 0.25–0.89; p = 0.021). Even modest reductions in pH were associated with significantly higher mortality risk. Acid–base disturbances in COVID-19 may reflect renal dysfunction, tissue hypoperfusion, lactic acidosis, or metabolic decompensation [20–22]. Acidemia may further impair cardiovascular function and hemodynamic stability [23]. The persistence of this association after adjustment for renal function and inflammatory markers suggests that acid–base status provides integrative prognostic information beyond isolated laboratory parameters.

Serum creatinine independently predicted mortality (adjusted OR 1.37; 95% CI 1.05–1.77; p = 0.019) but did not retain statistical significance in the composite outcome model (p = 0.069). This difference highlights the importance of analyzing mortality separately from composite endpoints. Renal dysfunction may reflect systemic organ failure and reduced physiological reserve, contributing more strongly to fatal outcomes than to respiratory failure alone.

Chest CT findings were analyzed in univariable models. Although morphological patterns were common—particularly ground-glass opacities, bilateral involvement, multifocal, and peripheral lesions—the extent of pulmonary involvement was the most relevant prognostic parameter. Parenchymal involvement exceeding 50% was associated with nearly threefold higher odds of death (OR 2.87; 95% CI 1.42–5.79; p = 0.003). This supports previous evidence that quantitative assessment of disease burden may be more prognostically informative than descriptive radiological patterns [24,25]. CT variables were not included in multivariable models due to incomplete availability and potential collinearity with clinical severity markers.

The composite outcome of death or invasive mechanical ventilation yielded largely consistent results, with four of five predictors remaining significant. The lower explanatory power of this model (Nagelkerke R² = 0.34) likely reflects the heterogeneity inherent in composite endpoints. The consistency of age, NLR, comorbidity burden, and arterial pH across both models reinforces their robustness as early prognostic markers.

Methodologically, this study adopted an a priori variable selection strategy based on clinical relevance and pathophysiological representation, avoiding automated stepwise procedures and respecting events-per-variable principles. Complete-case analyses were performed without imputation, prioritizing variables with high data availability to preserve sample size.

### Strengths and Limitations

Strengths include the comprehensive characterization of 76 admission variables, high availability of arterial blood gas data, standardized CT interpretation, and representation of an underreported region of Brazil. Limitations include the retrospective design, limited number of events (45 deaths), missing data for certain biomarkers, absence of longitudinal laboratory trends, and lack of post-discharge follow-up. CT variables were analyzed only in univariable models. As a single-center pre-vaccination cohort, external validation is required before extrapolation to contemporary populations.

## Conclusion

In this retrospective cohort of 482 patients hospitalized with COVID-19 during the pre-vaccination era, five admission variables—age, arterial pH, neutrophil-to-lymphocyte ratio, comorbidity burden, and serum creatinine—were independently associated with in-hospital mortality. These routinely available clinical and laboratory parameters represent distinct pathophysiological domains and collectively explained a substantial proportion of mortality variance. Arterial pH provided independent prognostic information beyond inflammatory and renal markers. These findings support early risk stratification using readily obtainable admission data and provide a reference framework for future validation studies.

## Data Availability

All relevant data are within the paper and its Supporting Information files.

## Acknowledgments

The authors gratefully acknowledge the nursing staff, respiratory therapists, laboratory technicians, and all healthcare workers at Hospital do Coração Anis Rassi who provided dedicated patient care during the COVID-19 pandemic. We also thank the administrative and data management teams for their support in medical record retrieval and data collection. This work is dedicated to the memory of Professor Anis Rassi (1929–2021), founder of Hospital do Coração Anis Rassi, whose vision of excellence in patient care made this institution — and this study — possible.

## Author contributions

**Conceptualization:** ARJ, VMR.

**Methodology:** ARJ, VMR, GFOB.

**Software:** ARJ.

**Validation:** ARJ, VMR, GFOB, JVRG.

**Formal analysis:** ARJ.

**Investigation:** ARJ, VMR, HMG, EPS, JVRG, GPC, GFOB, CRK, FMS, FMR, VRPC, AFC, GGR.

**Resources:** RNDRF.

**Data curation:** ARJ.

**Writing – original draft:** ARJ.

**Writing – review & editing:** ARJ, VMR, JVRG, HMG, CRK, FMS, GFOB, EPS, FMR, GPC, VRPC, RNDRF, AFC, GGR.

**Visualization:** ARJ, VMR.

**Supervision:** ARJ, RNDRF.

**Project administration:** ARJ.

## Supporting Information

S1 Table. In-hospital complications according to mortality.

S2 Table. Admission location, length of stay, medications, oxygen therapy, and treatment interventions.

S1 Fig. Prevalence of comorbidities and risk factors among 482 hospitalized COVID-19 patients.

S2 Fig. Distribution of time from symptom onset to hospital admission. S3 Fig. Frequency of symptoms at hospital admission.

S4 Fig. Disease severity classification according to WHO and NIH criteria. S5 Fig. Complications during hospitalization.

S6 Fig. Distribution of length of hospital stay. S7 Fig. Medications used during hospitalization.

S8 Fig. Oxygen and ventilatory support during hospitalization.

S9 Fig. Forest plots of univariate logistic regression analysis for predictors of in-hospital mortality.

